# Visual Scoring of Chest CT at Hospital Admission Predicts Hospitalization Time and Intensive Care Admission in Covid-19

**DOI:** 10.1101/2020.10.30.20222471

**Authors:** Erik Ahlstrand, Sara Cajander, Per Cajander, Edvin Ingberg, Erika Löf, Matthias Wegener, Mats Lidén

**Affiliations:** Department of Medicine, Faculty of Medicine and Health, Örebro University, S-701 82 Sweden; Department of Infectious Diseases, Faculty of Medicine and Health, Örebro University, S-701 82 Sweden; Department of Anesthesiology and Intensive Care, Faculty of Medicine and Health, Örebro University, S-701 82 Sweden; Department of Infectious Diseases, Örebro University Hospital, Region Örebro County, PO Box 1613, S-701 16 Örebro, Sweden; Department of Radiology, Örebro University Hospital, Region Örebro County, PO Box 1613, S-701 16 Örebro, Sweden; Department of Radiology, Faculty of Medicine and Health, Örebro University, S-701 82 Sweden

## Abstract

**Background:** The extent and character of lung involvement on chest computerized tomography (CT) have a prognostic value in covid-19 but there is lack of consensus on how to assess and stage CT features. A scoring system of lung involvement in covid-19, Örebro covid-19 Scale (ÖCoS) was implemented in clinical routine on April 1 2020 in Örebro Region, Sweden. The ÖCoS-severity score measures the extent of lung involvement while ÖCoS-temporal stage characterizes the parenchymal involvement. The objective of the present study was to evaluate the ÖCoS scores in relation to clinical outcome of covid-19.

**Methods:** Population based study including data from all hospitalized patients with covid-19 in Örebro Region during March to July 2020. Chest CT scores at the time of hospital admission and ICU admission were analyzed in relation to hospital and intensive care unit (ICU) length of stay, time to ICU admission and admission to ICU or death.

**Findings:** In the 381 included patients, there was a close correlation of the ÖCoS-severity score on admittance to hospital and the hospital length of stay. The ÖCoS-severity score on hospital admittance was a strong predictor for both a severe outcome in regards to ICU admittance or death and the time to ICU admittance. On admittance to ICU, both ÖCoS-severity score and temporal stage were correlated with the ICU length of stay.

**Interpretation:** Chest CT visual scoring on admission to hospital predicts the clinical course in covid-19 pneumonia.

**Funding:** This work was supported by the Örebro Region, Sweden.

## Introduction

The novel coronavirus disease (covid-19), caused by the severe acute respiratory syndrome coronavirus 2 (SARS-CoV-2), is a global pandemic that represents an important threat to human health. Up to September 28, 2020, more than one million people have died from covid-19 in over 160 countries^1^. Although most covid-19 patients present with mild illness, a minority of patients have a severe outcome characterized by pneumonia, respiratory failure and acute respiratory distress syndrome (ARDS)^2^. The strongest independent risk factor for a severe outcome is high age^3^. Other important risk factors include male sex, cardiovascular disease, and diabetes with complications^3,4.^ Within certain populations up to 30% of infected patients may require hospitalization and increased medical support^5^. Outbreaks of covid-19 are causing a considerable strain on the health system with a shortage of hospital and intensive care units (ICU) beds^6^. To manage the potentially critical burden on the health care system during outbreaks and to triage individual patients, there is a clinical need for robust prognostic models to predict the course of covid-19.

Previous research have demonstrated a prognostic role of the extent and character of lung involvement on chest computerized tomography (CT) in covid-19^7-9^. These studies have applied an abundance of different methods to measure covid-19 lung involvement and consequently there is no consensus in the literature on how to assess and stage CT features of covid-19. In addition, published predictive models have generally been developed using retrospectively interpreted CT images by expert thoracic radiologists in contrast to clinical routine were chest CTs typically are read by general radiologists.

In response to an increased demand for chest CTs in covid-19, a concise scoring system of lung involvement in covid-19, the Örebro covid-19 Scale (ÖCoS) was implemented in clinical routine on April 1 2020 at the Department of Radiology, Örebro Region, Sweden. The intention was to provide a standardized assessment of covid-19 pneumonia. Both the extent of lung involvement, ÖCoS-severity score, and the character of involvement, ÖCoS-temporal stage, are assessed on the scale.

The current study is a population-based evaluation of the clinically provided ÖCoS chest CT scores, including all patients hospitalized for covid-19 in the Örebro Region during the first five months, March-July 2020, of the covid-19 outbreak in Sweden. The primary aim was to evaluate chest CT at hospital admission as a predictor of hospital length of stay (LoS) and admission to ICU or mortality. The secondary aim was to evaluate chest CT at ICU admission in relation to ICU LoS.

## Material and Methods

### Ethics

The Swedish Ethical Review Authority approved the study protocol and waived the informed consent requirement for this retrospective study, reference number 2020-02515.

### Study population

The study included all patients ≥18 years admitted as in-patients due to laboratory confirmed covid-19 in one of the three hospitals, one university hospital and two associated hospitals, in Örebro Region, Sweden. Covid-19 patients were identified through the ICD-codes corresponding to either a primary laboratory confirmed diagnosis of covid-19, or a primary diagnosis of covid-19 based on a typical clinical picture in combination with a positive antibody test for covid-19, or a laboratory confirmed secondary diagnosis of covid-19 with a non-etiological pulmonary diagnosis as a primary diagnosis.

### Data source

Data regarding age, sex, hospitalization times, hospitalization routes, ICU admission, death during and after hospitalization, laboratory tests for covid-19, and radiology reports were extracted from the information system and radiological information system of the Örebro Region. Data from March 1 to August 31, 2020 were extracted, but only patients admitted to hospital before July 4 were included to enable at least 60 days observation time in the extracted data. Patients arriving from hospitals external from the Örebro Region were excluded. Figure 1 describes the inclusion process in detail.

**Figure 1.**
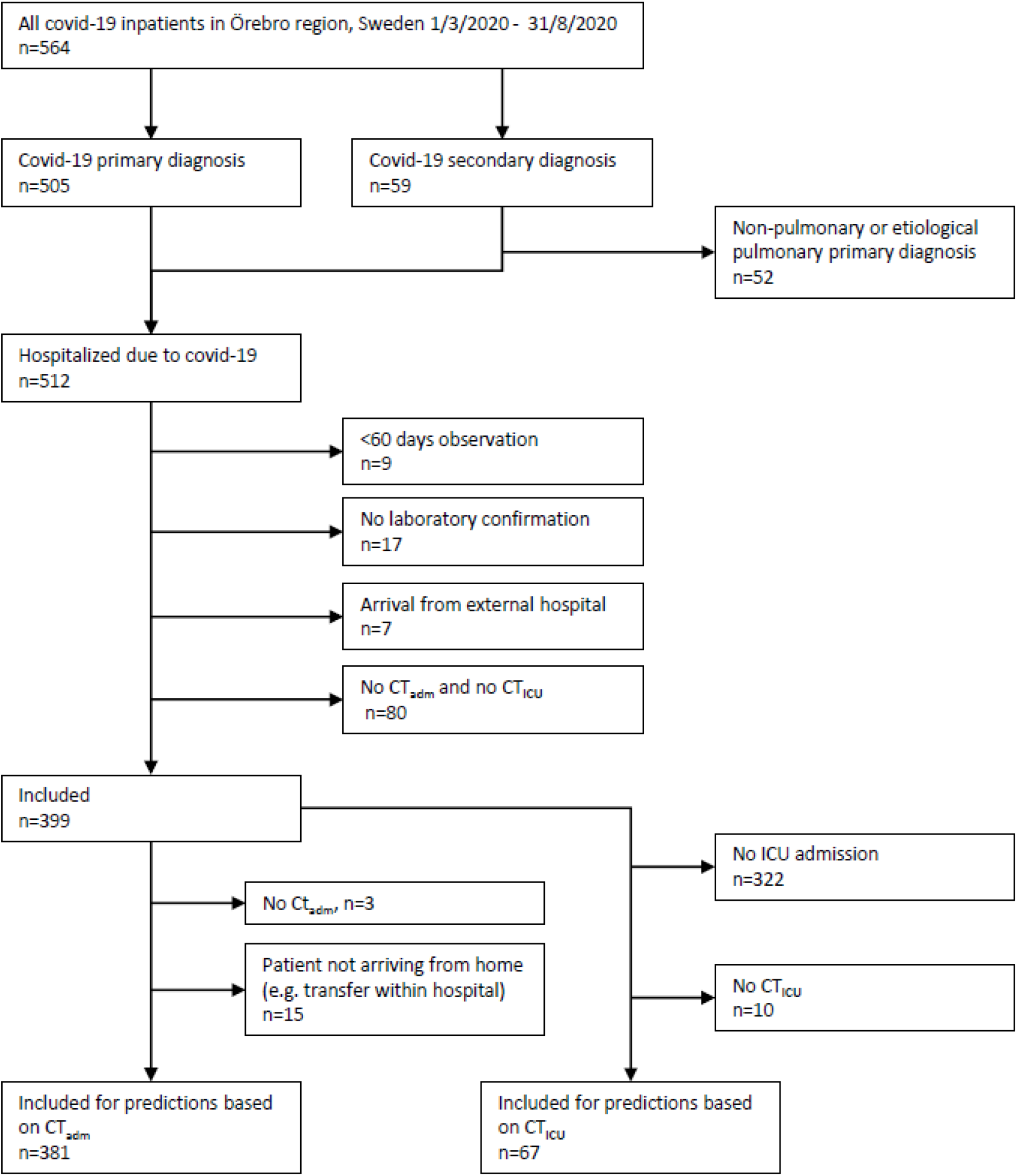
Inclusion and exclusion flowchart. Note: ICU – Intensive Care Unit. CT_adm_ – CT at hospital admission. CT_ICU_ – CT at ICU admission.

### Chest CT

#### Visual scoring – Örebro Covid-19 Scale (ÖCoS)

The structured ÖCoS chest CT report was introduced on April 1, 2020. The scale consists of the disease severity score (ÖCoS-severity score) and temporal stage (ÖCoS-temporal stage) on discrete scales, Figure 2. The ÖCoS-severity score is a visual assessment of the global lung involvement on a six-point scale (0%, <10%, 10-25%, 25-50%, 50-75%, >75%) whereas the ÖCoS-temporal score is a five-point ordinal scale assessing the parenchymal characteristics based on the transition from normal parenchyma, via ground glass opacities (GGO) to consolidations as described in early reports of the covid-19 disease evolution^10^. Radiologists were instructed to provide only one selection for temporal stage and one selection for severity score for each examination. Scores were provided similarly regardless of whether covid-19 was confirmed or not confirmed at the time of reading. Figure 3 gives examples of ÖCoS scores.

**Figure 2.**
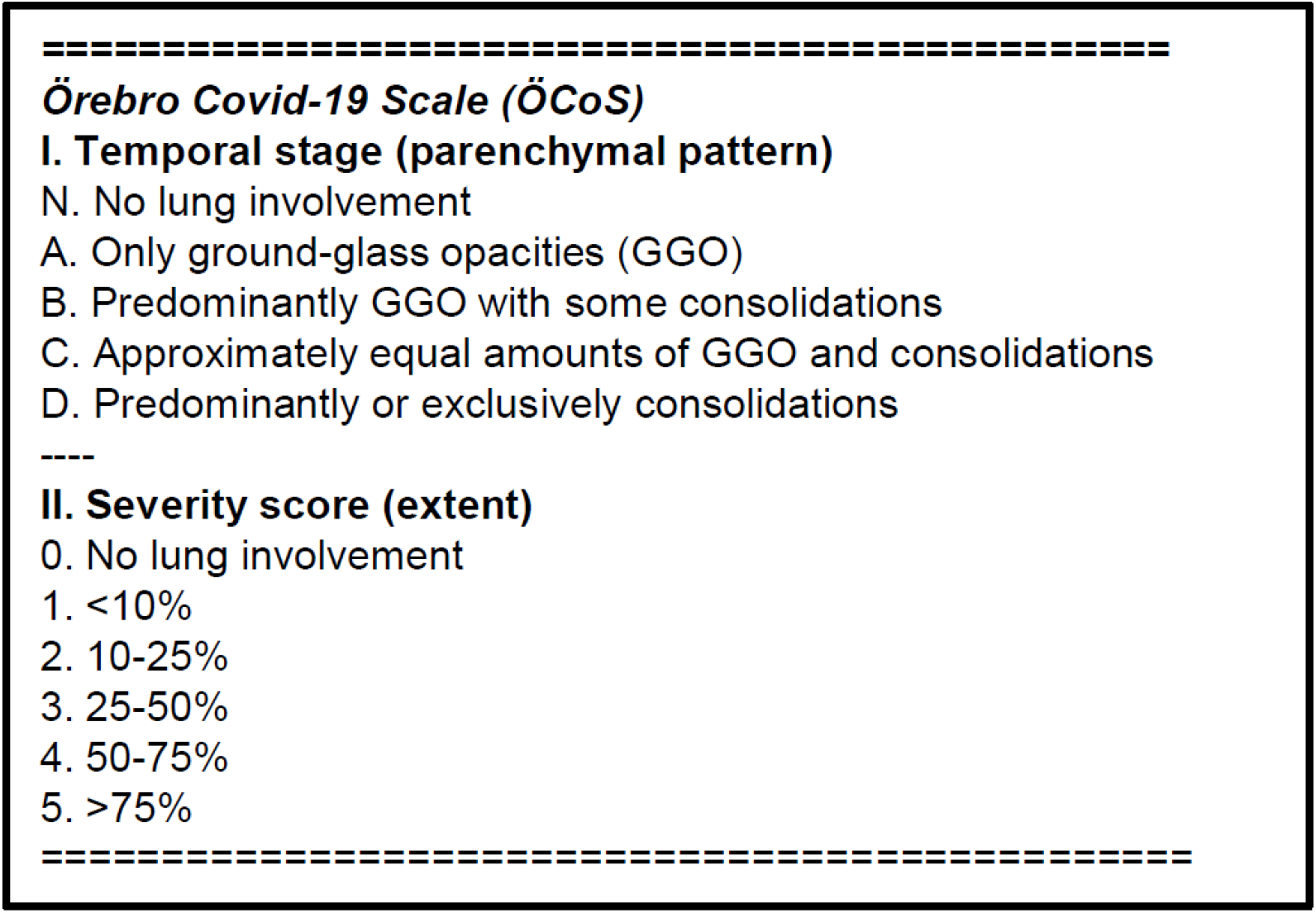
The Örebro Covid-19 Scale. GGO – ground glass opacities. Crazy-paving pattern was assessed as GGO and organizing pneumonia pattern as consolidations. Only one selection for temporal stage and one selection for severity score was allowed. Stage N was always combined with severity 0-(N/0), and stage A-D was always combined with severity 1-5.

**Figure 3.**
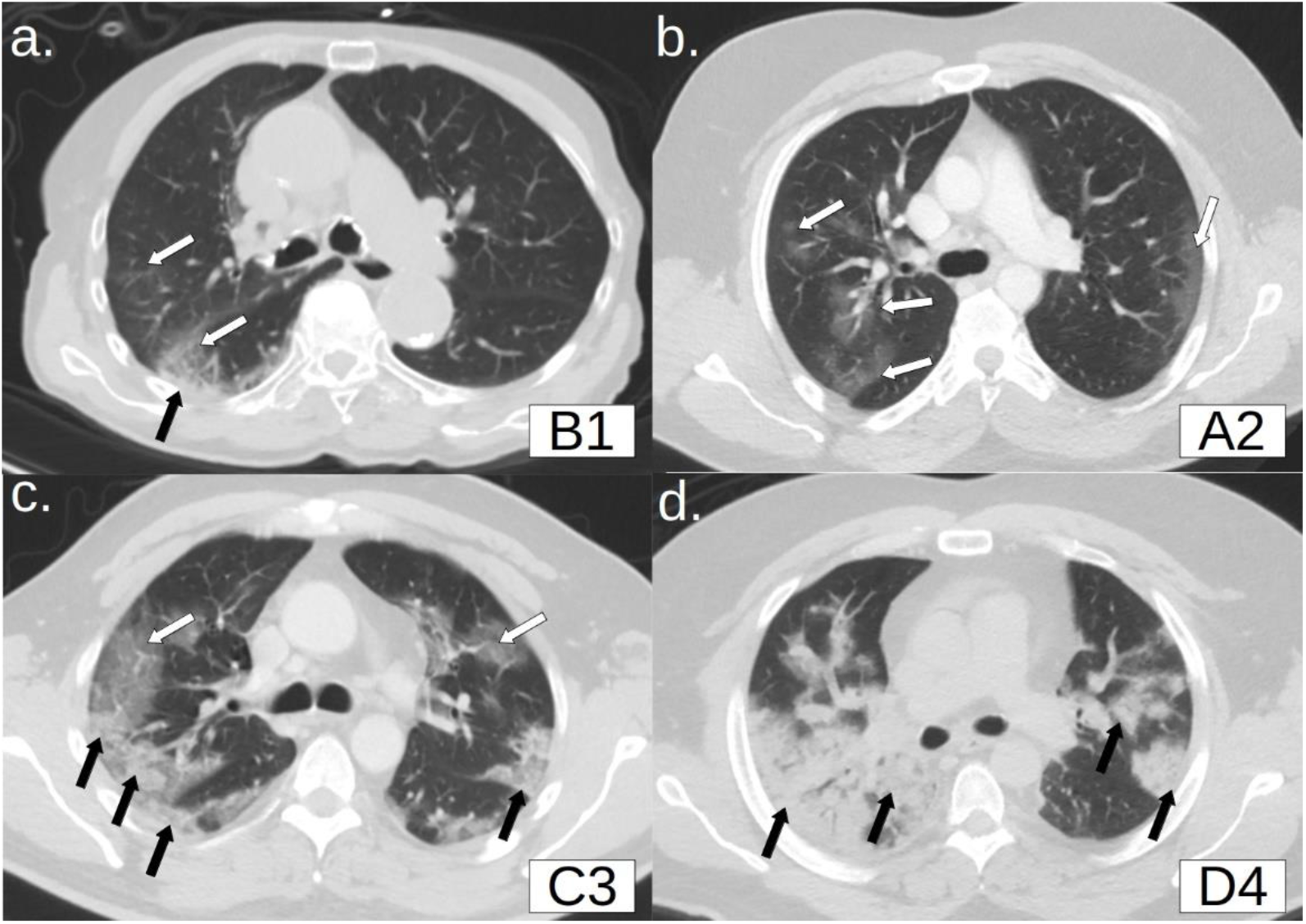
Lung window chest CT axial images at the level of carina demonstrating examples of Örebro Covid-19 Scale (ÖCoS) temporal stage and severity score. White arrows – ground-glass opacities (GGO). Black arrows – consolidations. a. ÖCoS-temporal/severity B/1 (predominantly GGO, <10% lung involvement). b. ÖCoS A/2 (Only GGO, 10-25% lung involvement). c. ÖCoS C/3 (Approximately equal GGO and consolidations, 25-50%). d. ÖCoS D/4 (Predominantly consolidations, 50-75%).

In the study, the ÖCoS scores were extracted from the clinical radiology reports. Approximately 30 different radiologists and residents provided clinical scores that were extracted for the study. In cases where no clinical ÖCoS scores were provided, mostly because of nighttime overseas teleradiological reading and CT preformed before April 1, a separate ÖCoS reading for the study was performed by a radiology resident (MW) blinded to all clinical information.

#### CT timing

The CT at hospital admission (CT_adm_) was defined as the chest CT closest in time to hospital admission, with no longer than two days difference. The CT at ICU transfer (CT_ICU_) was defined as the chest CT closest in time to ICU transfer, with no longer than two days difference.

### Nucleic acid amplification and antibody tests

For detection of SARS-CoV-2 RNA, nasopharyngeal swab specimens were analyzed by different methods during the study period. The vast majority of samples were analyzed by an in-house real-time reverse-transcription polymerase chain reaction (RT-PCR) targeting the E gene (with an RdRp gene assay as confirmation) adapted from the protocol recommended by WHO, or the RdRp gene assay alone. For antibody testing, the Diasorin (Saluggia, Italy) Liaison XL test for SARS-CoV-2 IgG was used, in combination with Euroimmun (Lübeck, Germany) SARS-CoV-2 IgG ELISA for confirmation in weakly positive samples to increase specificity.

### Outcome measures

#### Hospital length of stay

To summarize the effect of covid-19 on hospitalization time in the presence of the competing event of death, we used the composite measure hospital free days 60 days post-admission (HFD_60_). For each patient, the total number of HFD_60_, including readmissions, during the 60 days following the first admission to hospital with covid-19 was computed. The hospital length of stay (LoS) was defined as 60-HFD_60_. This outcome equals the hospitalization time within 60 days in non-deceased patients whereas deceased patients and patients with a hospitalization time over 60 days will have a hospital LoS of 60.

#### Combined ICU admission and mortality rate

The combined risk for ICU admission or death within 60 days was used as outcome measure in a multivariate logistic regression.

#### Time to ICU admission

The intervals in days between CT_adm_ and ICU admission were derived for all patients admitted to an ICU.

#### ICU length of stay

For patients admitted to an ICU, the 60-day ICU free time (IFD_60_) following the day of ICU transfer was computed. The ICU LoS used in the analysis was 60-IFD_60_, which corresponds to the total ICU-time within 60 days in non-deceased patients.

### Statistics

Matlab R2020a (The MathWorks Inc., Natick, MA) was used for statistics.

Multivariate linear regression with 60-HFD_60_ as dependent variable was performed to identify the predictors for LoS. Age was treated as a continuous variable whereas ÖCoS temporal stage, ÖCoS severity score and sex were treated as categorical variables. A reduced model was developed, where the temporal stage A, B and C were grouped, forming the temporal stages: N (No lung involvement), ABC (GGO extent greater than or equal consolidation extent), and D (Predominantly consolidations), Figure 2. Only linear terms with no interactions were included in the models. Twenty-fold cross validation was performed to assess overfitting on the reduced linear regression model with LoS as dependent variable.

For the analysis of time to ICU admission, Spearman correlation coefficient was computed for CT_adm_ ÖCoS-severity score, CT_adm_ ÖCoS-temporal stage and age, and Wilcoxon signed rank test was used to assess dependency of patient sex.

For the analysis of ICU length of stay, Spearman correlation coefficient was used to assess dependency of CT_ICU_ ÖCoS-severity score, CT_ICU_ ÖCoS-temporal stage and age, and Wilcoxon signed rank test was used to assess dependency of patient sex.

### Funding

This work was supported by the Örebro Region, Sweden.

## Results

### Patient characteristics

In 375 out of 381 included patients, SARS-CoV-2 RNA was confirmed by RT-PCR and the additional six patients were included on basis on a typical clinical picture supported by a positive covid-19 antibody test. Patient characteristics and outcomes are described in Table 1.

**Table 1.** Patient demographics, outcomes and ÖCoS scores at hospital and intensive care unit admission.

|  | **Total cohort**<br>**Count (%)** | **ICU cohort**<br>**Count (%)** |
| --- | --- | --- |
| **Cohort size** | 381 | 67 |
| **Sex, female** | 184 (48.3%) | 23 (34.3%) |
| **Age, years\*** | 60 (50-74) | 58 (50-68) |
| **>70 years** | 120 (31.5%) | 10 (14.9%) |
| **Hospital free days**<br>**within 60 days\*** | 51 (39-56) | 43 (20-49) |
| **Admitted to ICU** | 74 (19.4%) | .. |
| **Death** | 46 (12.1%) | 10 (14.9%) |
| **ÖCoS-temporal stage** | **at hospital**<br>**admission** | **at ICU**<br>**admission** |
| **Typ N (No lung**<br>**involvement)** | 25 (6.6%) | 0 (0.0%) |
| **Typ A (Only GGO)** | 84 (22.0%) | 7 (10.4%) |
| **Typ B (GGO with**<br>**some consolidations)** | 125 (32.8%) | 21 (31.3%) |
| **Typ C (equal**<br>**amounts of GGO and**<br>**consolidations)** | 56 (14.7%) | 17 (25.4%) |
| **Typ D**<br>**(Predominantly or**<br>**exclusively**<br>**consolidations)** | 91 (23.9%) | 22 (32.8%) |
| **ÖCoS-severity score** | **at hospital**<br>**admission** | **at ICU**<br>**admission** |
| **0 . No lung**<br>**involvement** | 25 (6.6%) | 0 (0.0%) |
| **1 <10%** | 86 (22.6%) | 2 (3.0%) |
| **2 10-25%** | 135 (35.4%) | 6 (9.0%) |
| **3 25-50%** | 95 (24.9%) | 28 (41.8%) |
| **4 50-75%** | 33 (8.7%) | 25 (37.3%) |
| **5 >75%** | 7 (1.8%) | 6 (9.0%) |

Note: \* Median (interquartile range)

Clinically provided ÖCoS scores were available in 309 out of 381 CT_adm_, and in 53 out of 67 CT_ICU_.

During the retrospective inclusion period, in-patient covid-19 patients were treated with a standard of care in line with international recommendations of that time^11,12^, including oxygen support and low-molecular weight heparins.

### Hospital length of stay

The hospital length of stay (LoS) in relation to CT_adm_ ÖCoS-severity scores are shown for different age groups in Figure 4. In patients ≤70 years old there was a close correlation between the ÖCoS severity score and the LoS, while the ÖCoS-severity score was less clearly correlated to the LoS in older patients.

**Figure 4.**
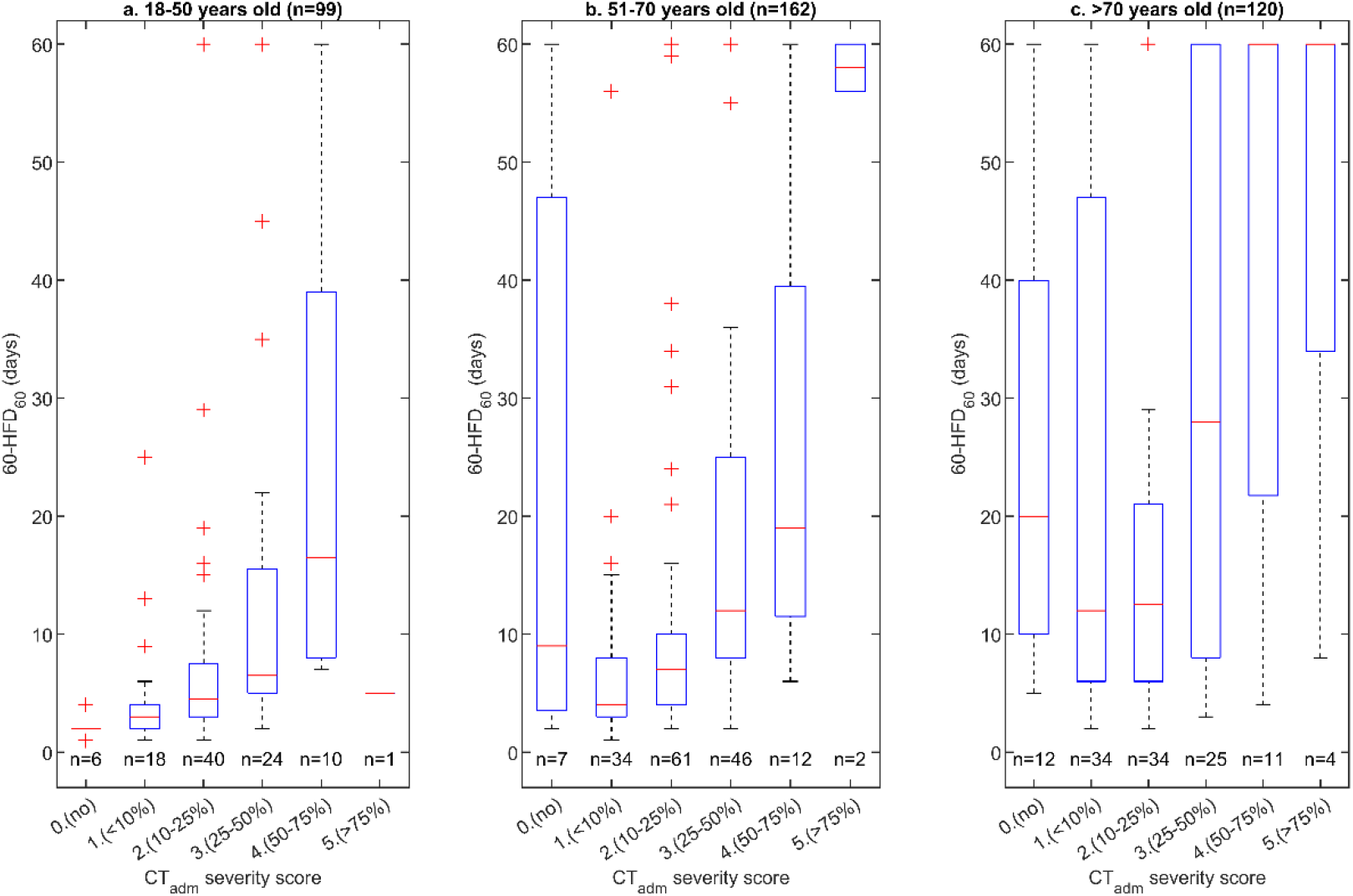
Box plot showing hospital length of stay, defined as 60-HFD_60_, in relation to ÖCoS severity score on CT at hospital admission for a) 18-50 years old, b) 51-70 years old, and c) >70 years old. HFD_60_ – 60-day Hospital Free Living Days

The multivariate regression analysis identified patient sex, age and ÖCoS-severity score as statistically significant predictor variables for LoS, Table 2. Since temporal stage A, B and C demonstrated similar coefficients in the multivariate analysis, a reduced model was developed with temporal stage A, B and C grouped. In the reduced model, the temporal stage was a statistically significant predictor variable, and there was a consistent reduction in hospital LoS for temporal stage D (predominantly consolidations) compared to earlier stages A-C (demonstrating more GGO) and the first stage N (no lung involvement), Table 2.

**Table 2.** Multivariate linear regression with hospital length of stay, defined as 60-HFD_60_ as dependent variable. n=381.

|  | Multivariate model |  | Reduced model |  |
| --- | --- | --- | --- | --- |
|  | Coefficient (days) | p-value | Coefficient (days) | p-value |
| Constant term | -7.3 | 0.13 | -7.3 | 0.13 |
| Age | 0.40 | <0.001 | 0.40 | <0.001 |
| Sex (M vs. F) | 4.5 | 0.012 | 4.5 | 0.013 |
| CT temporal stage |  | 0.16 |  | 0.048 |
| - N. No lung involvement | Reference |  | Reference |  |
| - A. GGO exclusively | -5.9 | 0.15 |  |  |
| - B. GGO > consolidation | -7.7 | 0.067 | -6.6 (A/B/C) | 0.092 (A/B/C) |
| - C. GGO ≈ consolidation | -6.5 | 0.16 | | |
| - D. predominantly or exclusively consolidations | -10.4 | 0.017 | -10.1 | 0.018 |
| CT severity score |  | <0.001 |  | <0.001 |
| - 0-1. <10% | Reference |  | Reference |  |
| - 2. 10%-25% | 1.4 | 0.56 | 1.2 | 0.62 |
| - 3. 25% - 50% | 10.8 | <0.001 | 10.5 | <0.001 |
| - 4. 50% - 75% | 20.3 | <0.001 | 20.0 | <0.001 |
| - 5. >75% | 30.2 | <0.001 | 29.7 | <0.001 |

Note: GGO – ground-glass opacities. HFD<sub>60</sub> – 60-day hospital free days.

The root mean square errors (RMSE) of the full model and the reduced model were similar, 17.2 days, indicating little loss of information in the reduction of predictors. Twenty-fold cross validation of the reduced model linear regression showed a comparable RMSE, 17.6 days, indicating only minor overfitting in the model.

The coefficients in the linear regression provides an interpretation of the impact of each variable in terms of LoS days: the LoS increases with four days per ten years age difference and with four days in males compared to females. A higher ÖCoS severity score is associated with longer LoS: Compared to ÖCoS 0-1 (<10% extent), LoS in patients with CT_adm_ demonstrating ÖCoS 2 (10-25%) increased one day, ÖCoS 3 (25-50%) eleven days, ÖCoS 4 (50-75%) 20 days, and ÖCoS 50020(>75%) 30 days.

A more advanced ÖCoS-temporal stage, suggesting a later phase of covid-19 pneumonia at hospital admission, was associated with a shorter LoS. Compared to ÖCoS N (no lung involvement), LoS in patients with CT_adm_ demonstrating ÖCoS A-C (GGO extent up to equal consolidation extent) decreased seven days and ÖCoS D (predominantly consolidations) decreased ten days.

### ICU admission and mortality rate

Figure 5 shows the combined ICU admission and death rate in relation to ÖCoS-severity score at hospital admission. In the multivariate logistic regression analysis, patient age was dichotomized as over or under 70 years. The analysis identified the ÖCoS-severity score at hospital admission, *p*<0.001, patient sex, *p*=0.006 and age, *p*=0.002 as significant predictor variables for the combined outcome of ICU admission and mortality, Table 3.

**Table 3.** Logistic regression with outcome Intensive Care Unit (ICU) admission or death. n=381

|  | Multivariate model |  |
| --- | --- | --- |
|  | Odds Ratio (95% C.I) | *p*-value |
| Age >70 years old | 2.3 (1.3-3.9) | 0.002 |
| Sex (M vs. F) | 2.1 (1.2-3.4) | 0.006 |
| CT visual type |  | 0.91 |
| - N. No lung involvement | Reference |  |
| - A. GGO exclusively | 0.9 (0.3-3.2) | 0.93 |
| - B. GGO > consolidation | 0.7 (0.2-2.5) | 0.62 |
| - C. GGO ≈ consolidation | 0.7 (0.2-2.9) | 0.67 |
| - D. consolidation exclusively or predominantly | 0.7 (0.2-2.4) | 0.53 |
| CT visual extent |  |  |
| - 0-1. <10% | Reference |  |
| - 2. 10%-25% | 1.1 (0.5-2.3) | 0.84 |
| - 3. 25% - 50% | 4.4 (2.0-9.5) | <0.001 |
| - 4. 50% - 75% | 12.3 (4.6-33) | <0.001 |
| - 5. >75% | 12.1 (2.1-71) | <0.001 |

Note: GGO – ground-glass opacities.

**Figure 5.**
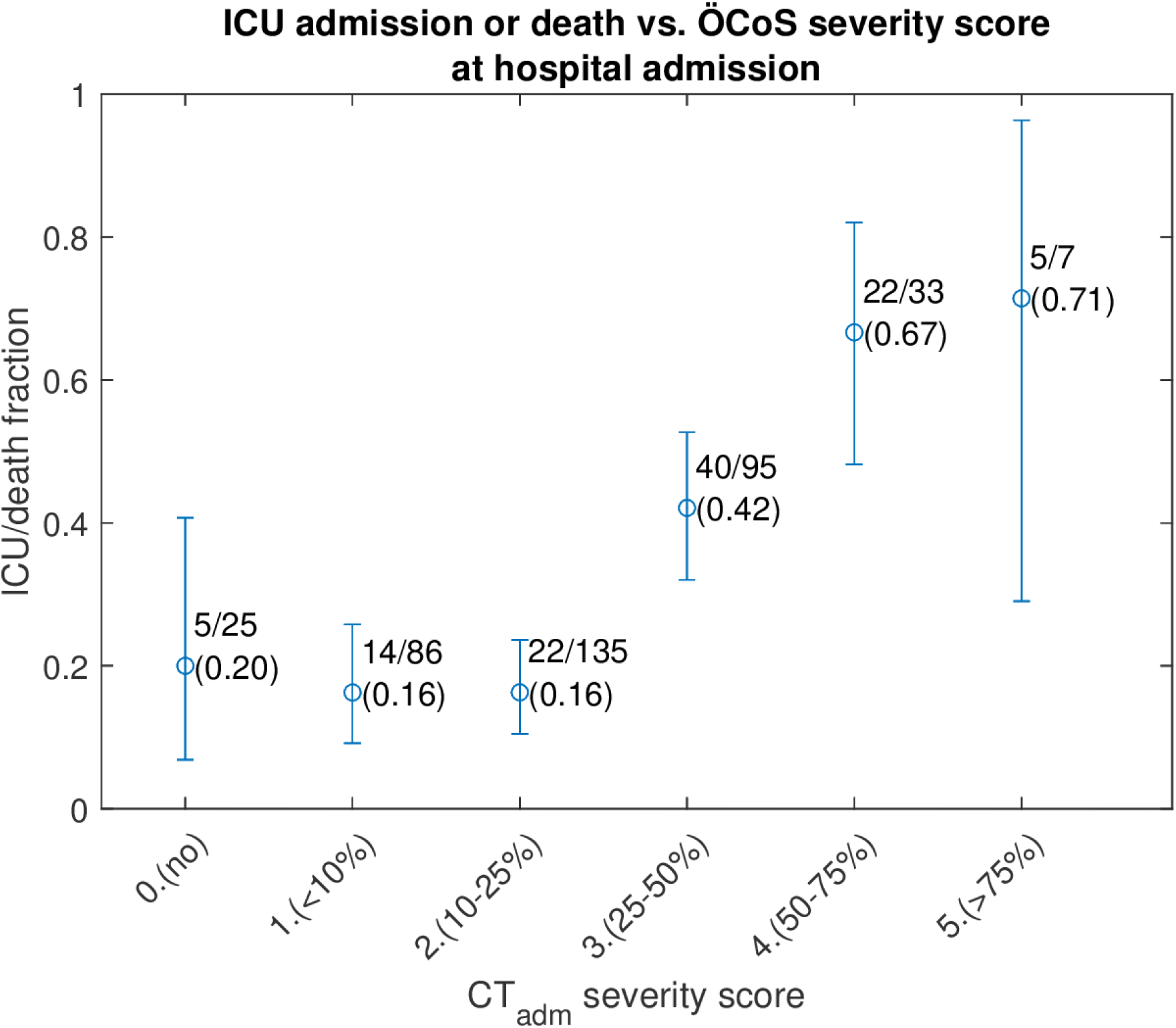
Combined ICU admission and 60-day mortality rate in relation to ÖCoS severity score on CT at hospital admission. Whiskers show 95% confidence interval for the proportion. ICU – Intensive Care Unit. ÖCoS – Örebro Covid-19 Scale.

### Time to ICU transfer

The interval between the CT_adm_ and ICU transfer was inversely related to the CT_adm_ ÖCoS-severity score, *p*=0.002, and CT_adm_ ÖCoS-temporal stage *p*=0.051, but was not significantly associated with age, *p*=0.15. There was no significant difference between male and female patients, *p*=0.39. The interval between CT_adm_ and ICU admission was longer for lower ÖCoS-severity scores and earlier ÖCoS-temporal stages, Figure 6.

**Figure 6.**
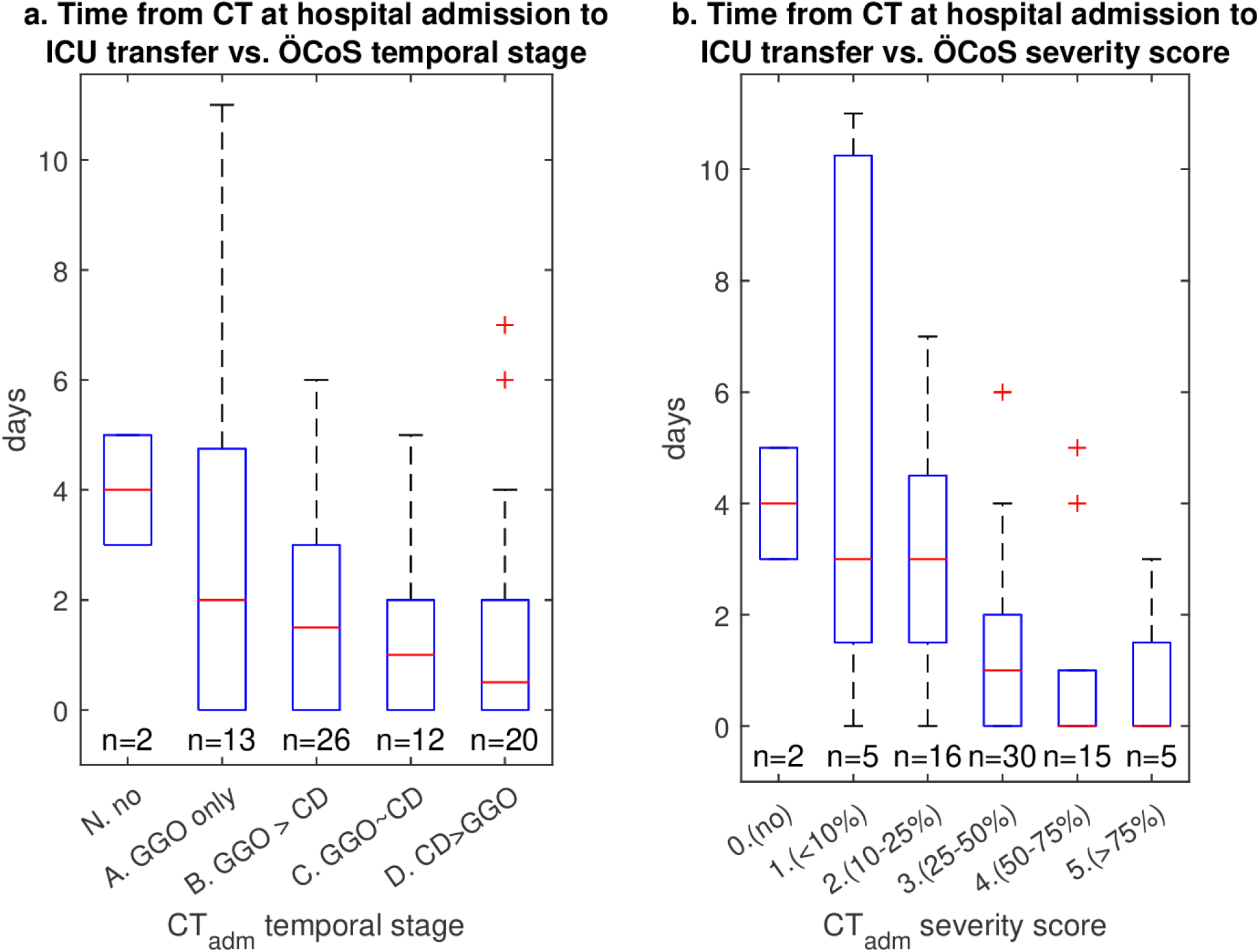
Time between first CT at hospital admission and ICU transfer according to (a) ÖCoS-temporal stage and (b) ÖCoS-severity score. ICU – Intensive Care Unit. ÖCoS – Örebro Covid-19 Scale

### ICU Length of Stay

The relation of ÖCoS scores on CT at the time of ICU transfer (CT_ICU_) and ICU outcomes are shown in Figure 7. The ICU LoS was positively correlated to CT_ICU_ ÖCoS-severity score, *p*<0.001, and inversely correlated to CT_ICU_ ÖCoS-temporal stage, *p*=0.044. The ICU LoS was correlated to patient age *p*<0.001, but not to patient sex *p*=0.33.

**Figure 7.**
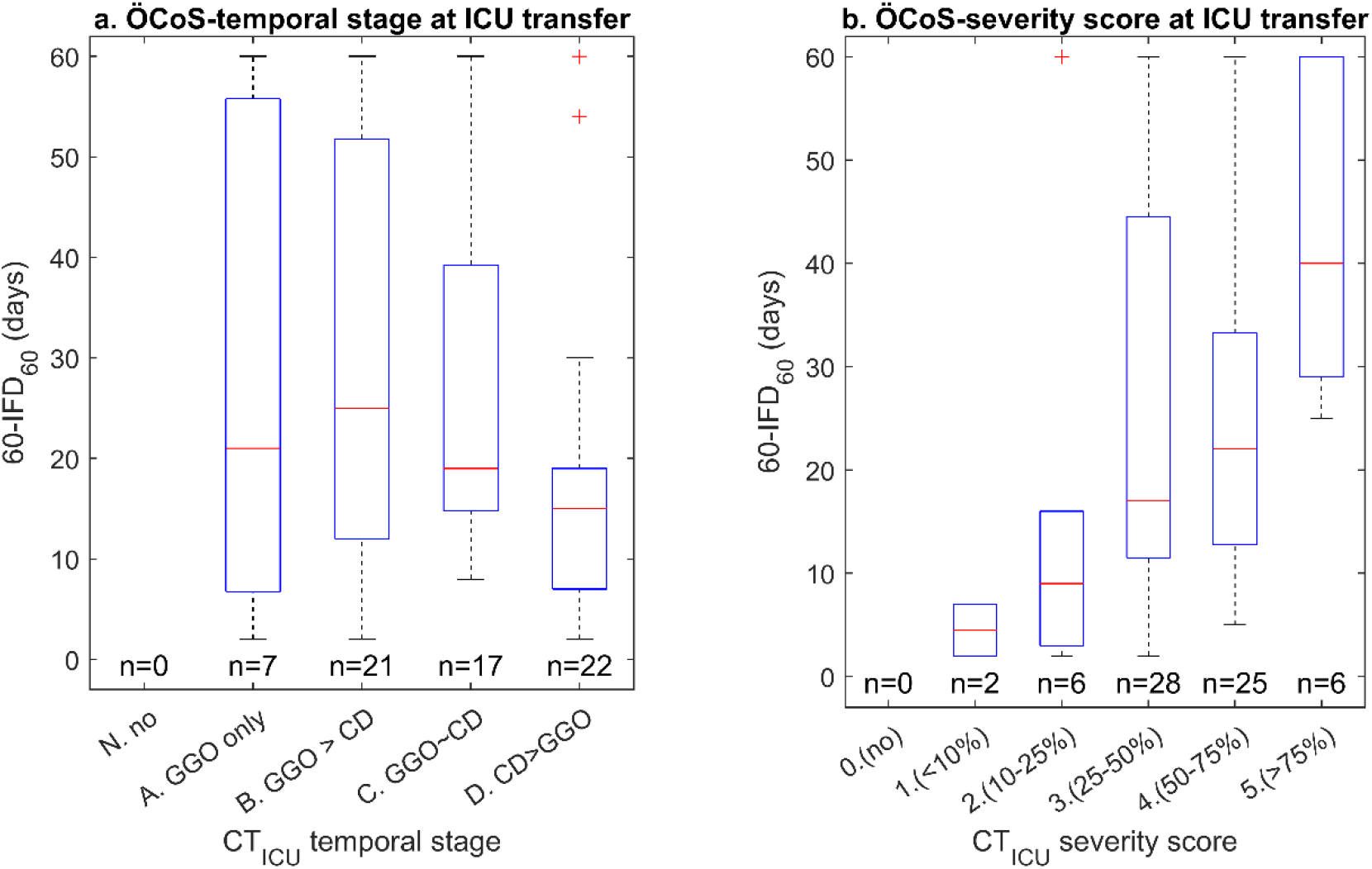
ICU length of stay, defined as 60-IFD_60_, in relation to (a) ÖCoS-temporal stage, and (b) ÖCoS-severity score in CT at ICU transfer. ICU – Intensive Care Unit. IFD_60_ = 60 day ICU Free Days. ÖCoS – Örebro Covid-19 Scale.

## Discussion

Covid-19 is an ongoing pandemic causing hospital crowding and shortage of ICU beds during outbreaks. The disease has a variable prognosis and established validated scores such as CURB-65 have a low overall performance^13,14^. Instead, we demonstrate that clinically provided chest CT visual scores at hospital admission can robustly predict the clinical course of covid-19 and that chest CT at ICU admission can predict ICU time, especially in patients up to 70 years old.

The two aspects of the ÖCoS visual score – the temporal development of the CT pattern from GGO to consolidations, and severity of lung involvement – are closely correlated to the patient outcomes in the present study. In particular, we demonstrate that the ÖCoS-severity score, a visual estimation of the extent of lung involvement, at hospital admission is a strong predictor of uneventful outcome in terms of death or ICU admission and that it is an important predictor for hospital LoS. The weaker association of these parameters in elderly patients, >70 years, may be due to frequent co-morbidities in the elderly group creating a more complex relationship.

Although different chest CT findings have been described in covid-19, the typical features are GGO and consolidations^15^. Three findings in the current study highlight that the transition from GGO to consolidation on the ÖCoS-temporal stage reflects the clinical course in the acute phase of covid-19, and often coincides with a deterioration of respiratory symptoms: 1) The inverse relationship of ÖCoS-temporal stage and time to ICU transfer, Figure 6, 2) the inverse relationship of ÖCoS-temporal stage at CT_ICU_ and ICU LoS, Figure 7 and 3) the lower number of hospital LoS days in late ÖCoS-temporal stages on CT_adm_, Table 2.

To put the current study in context, we performed a systematic literature search, Supplementary material. In summary, we found several reports on protocols of visual quantitative analysis of CT evaluated lung involvement demonstrating a correlation to the clinical severity of covid-19^9,16^. In addition, several semi-automatized^8,13,17^ and computerized^7,18,19^ quantitative measures of covid-19 lung involvement on CT have been described to be associated with outcomes related to a severe course of covid-19. However, to the best of our knowledge, up to date only one smaller study, published as a letter to the editor, reported real-life data on the predictive role of CT visual scoring in clinical routine^20^.

In this study, we report that the predictive role of chest CT can be reproduced in a non-selected population-based context with CT evaluations made by several reviewers as part of clinical routine. Since almost 80% of the study cohort underwent chest CT on admission there was probably only a limited degree of selection of patients referred for CT. We used a concise visual scoring system as a predictive model for outcome of hospitalized covid-19 patients. A strength is that the model apart from patient age and sex rely solely on CT findings, excluding clinical and laboratory data. The results indicate that triage with chest CT on admittance to hospital is a valuable tool for covid-19 patients, provided that a consistent scoring system is applied. The simplicity of the chest CT ÖCoS scoring enables a straightforward implementation in clinical practice, supported by its rapid acceptance among reading radiologists and referring clinicians in the Örebro Region, Sweden. Moreover, a strength of this study is that we could, in contrast to other studies, provide outcome-data up 60 days post-admission including mortality after hospital discharge.

The study has several limitations. Consistent with the inclusion criteria, the results only apply to in-patients. The use of clinically provided scores by multiple readers is a limitation, but also a strength in the study. Visual scoring is subjective and prone to interobserver variation, which reduces the precision of the provided scores. On the other hand, the scores used in the study are a reasonable estimate of the precision in a clinical scenario. Since the ÖCoS scores were provided in clinical routine, the reviewers were not formally blinded, but the main study outcomes of HFD_60_ and ICU admittance were naturally unknown to reviewers by the time of chest CT evaluation. Furthermore, we only included laboratory confirmed cases of covid-19, and consequently a few covid-19 cases are likely to have been excluded from the analysis^21^.

In conclusion, concise visual scoring of chest CT at hospital admission and at ICU transfer in clinical routine predicts the clinical outcome of covid-19, especially in patients <70 years. In situations where adjuvant treatments and hospital beds are limited, we believe that scoring of chest CT is informative and a valuable tool for clinical decision making.

## Supporting information

Tripod checklist

## Data Availability

Not available according to IRB decision.

## Supplementary material

A systemic literature search was performed on September 24th 2020 and 860 articles were retrieved from the Medline database via Pubmed using the search string (covid OR sars-cov-2) AND (CT OR “computed tomography” OR CAT OR “computer assisted tomography” OR “computerized tomography” OR “computerized tomography”) AND (prognos* OR predict* OR outcome). All titles were scanned for potential relevance and among the selected articles, abstracts were reviewed. Ultimately, the search resulted in 54 articles that were considered to address the question of interest, namely how chest CT images are associated with disease severity or outcome in patients with covid-19. A selection of relevant articles are referred to in the main text.

